# Immune Checkpoint Inhibitors as Independent and Synergistic Drivers of SJS/TEN: An Analysis of FAERS

**DOI:** 10.1101/2025.06.21.25330030

**Authors:** Eric Milan Mukherjee, Dodie Park, Amir Asiaee, Matthew S. Krantz, Cosby A. Stone, Michelle Martin-Pozo, Elizabeth Phillips

## Abstract

Immune checkpoint inhibitors (ICIs) are transformative cancer therapies but have been implicated in rare, life-threatening cutaneous adverse reactions such as Stevens-Johnson Syndrome and Toxic Epidermal Necrolysis (SJS/TEN). Whether ICIs serve as independent causative agents or amplify risk from other drugs remains incompletely understood.

**Methods:** We analyzed 13.9 million deduplicated reports from the FDA Adverse Event Reporting System (FAERS) between 2013 and 2023, including 17,495 cases of SJS/TEN. We conducted multivariable logistic regression to assess the independent effects of ICIs, high-risk (“strong”) and moderate-risk (“weak”) culprit drugs, as well as their interactions. Age was modeled using natural splines. Additive interaction was assessed using the relative excess risk due to interaction (RERI), attributable proportion (AP), and synergy index (S). We also performed Cox regression analyses using time-to-event (TTE) data to evaluate latency patterns associated with different ICI classes.

**Results:** ICI exposure was independently associated with increased risk of SJS/TEN (adjusted OR 9.14, 95% CI 8.42-9.93). Strong culprit drugs (e.g., allopurinol, TMP-SMX) and weak culprits (e.g., fluoroquinolones, macrolides) also conferred elevated risk. Notably, we observed significant additive interaction between ICIs and both strong (RERI 13.69, AP 0.38) and weak culprits (RERI 12.92, AP 0.52), indicating synergistic risk amplification. In time-to-event analyses of 4,086 cases with latency data, PD-1 inhibitors were consistently associated with delayed onset of SJS/TEN compared to non-ICI triggers (HR 0.71-0.82), with median latency nearly doubled.

**Conclusions:** Our findings support a two-hit immunopathogenic model in which ICIs lower the activation threshold for drug-specific T-cell responses, enabling otherwise tolerated medications to trigger severe cutaneous reactions. These results have critical implications for the co-prescription of high-risk drugs in patients receiving ICIs and underscore the need for enhanced pharmacovigilance and risk mitigation in cancer immunotherapy.

## Introduction

Immune checkpoint inhibitors (ICIs) are paradigm-shifting cancer treatments that are increasingly associated with Stevens-Johnson Syndrome, Toxic Epidermal Necrolysis (SJS/TEN) and other life-threatening cutaneous reactions. Differentiating ICI-induced “true” SJS/TEN from SJS/TEN-like reactions is difficult, the latter of which may be distinct lichenoid or bullous reactions.^1–3^ In some cases, ICI-related-SJS/TEN-like reactions occur in association with HLA restricted drug culprits like allopurinol, suggesting a “two-hit” mechanism.^4,5^ With increasing ICI use, a clearer understanding of their role in SJS/TEN is critical.

## Methods

We analyzed 13,986,839 deduplicated FAERS reports (2013-2023), containing 17,495 SJS/TEN cases. We assessed the impact of ICI using logistic regressions adjusted for age, sex, cancer, polypharmacy, strong (lamotrigine, TMP-SMX, phenytoin, allopurinol, carbamazepine) or weak (azithromycin, clarithromycin, erythromycin, ciprofloxacin, levofloxacin, moxifloxacin, and acyclovir) culprit exposure.

To assess latency patterns, we performed Cox proportional hazards analyses among SJS/TEN cases with documented latency. In Model 1, we used time-dependent Cox regressions with interval splitting to dynamically update exposure to PD-1, PD-L1, CTLA-4, or LAG-3 inhibitors. In Model 2, we compared latency between ICI-attributed and non-ICI-attributed cases, classifying primary suspect (PS) by ICI mechanism and using the same covariates.

## Results

In a multivariable logistic regression (**Figure 1**), ICI exposure was strongly associated with increased risk of SJS/TEN (adjusted OR [aOR]: 9.14, 95% CI: 8.42-9.93, p < 0.001). Strong culprit drugs were the most potent independent predictors (aOR: 14.31, 13.77-14.87). Interestingly, cancer diagnosis was inversely associated with SJS/TEN risk (aOR: 0.60, 0.58-0.63). Interaction terms revealed additive synergy between ICI exposure and culprit drugs. The ICI × strong culprit interaction yielded an attributable proportion (AP) of 0.38, indicating that 38% of the risk in co-exposed patients is attributable to interaction. For ICI × weak culprits, the AP was even higher (0.52).

**Figure 1.**
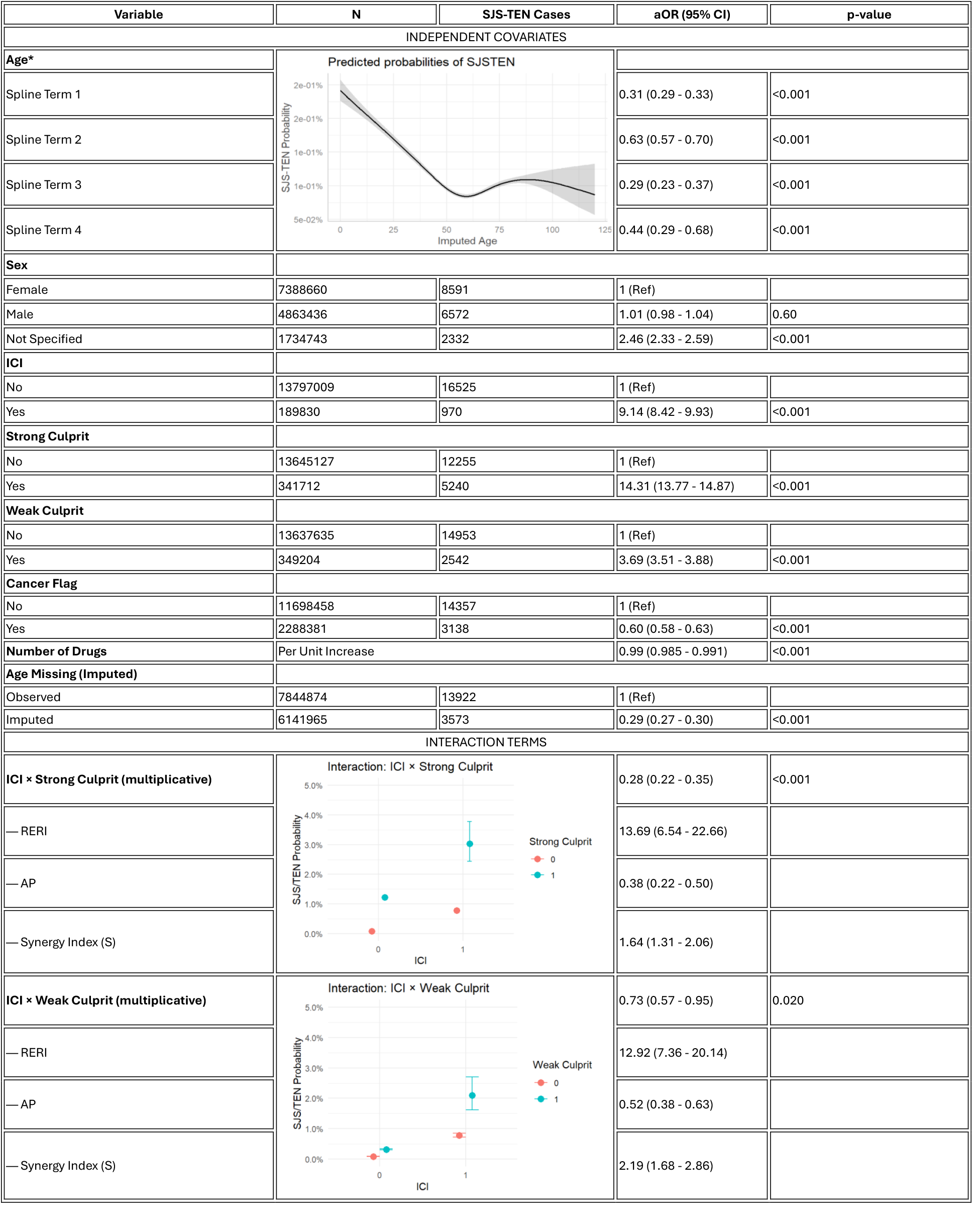
Multivariable logistic regression identifying independent and synergistic predictors of SJS/TEN. Adjusted odds ratios (aORs) with 95% confidence intervals are shown for independent covariates (age, sex, ICI use, strong and weak culprit drugs, cancer diagnosis, and polypharmacy) and for interaction terms between ICIs and culprit drug classes. Age was modeled using natural splines with 4 degrees of freedom. Additive interaction was quantified using the relative excess risk due to interaction (RERI), attributable proportion (AP), and synergy index (S). Notably, ICIs had a strong independent association with SJS/TEN (aOR 9.14) and exhibited substantial synergy with both strong and weak culprit drugs (RERI 13.69 and 12.92, respectively).

Patients with an anti-PD-1 as PS had TTE of 27 days, compared to 13 days for non-ICI, 15 days for PD-L1, and 20 days for CTLA-4/PD-1 combination. We further evaluated latency patterns using two Cox models. In the time-dependent exposure model/Model 1 (**Figure 2A**), PD-1 inhibitors were associated with delayed SJS/TEN onset (HR 0.82, *p* < .01). Patients with cancer diagnoses had later onset (HR 0.78, *p* < .001), as did those with greater polypharmacy (HR <1 per drug). In the causative-agent model/Model 2 (**Figure 2B**), PD-1 inhibitors again exhibited delayed onset compared to non-ICI causative drugs (HR 0.71, *p* < .001). The direction and magnitude of covariate effects were consistent between models.

**Figure 2.**
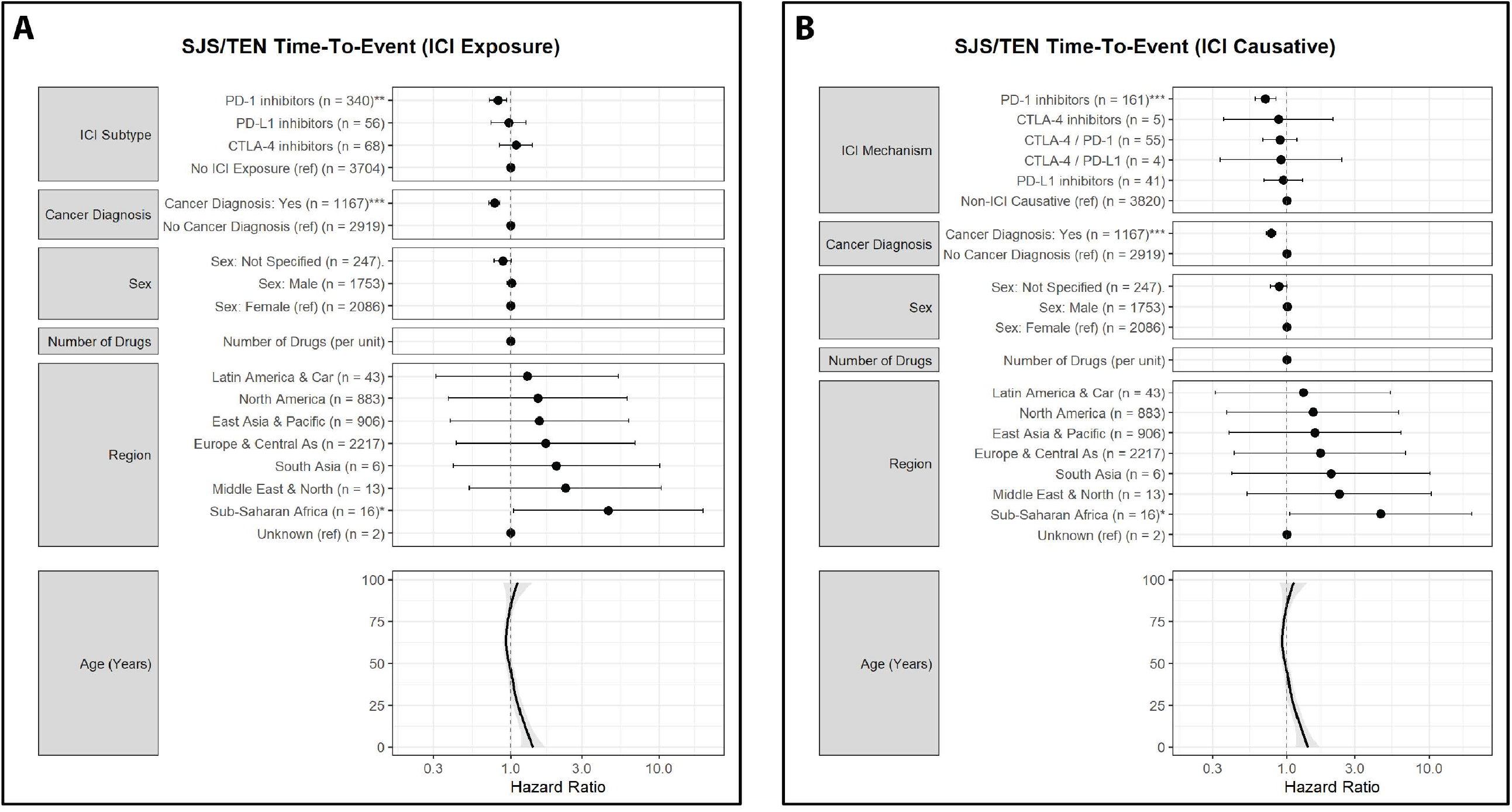
Latency Patterns of SJS/TEN in Relation to ICI Exposure and Mechanism of Causation. (A) Time-dependent Cox regression assessing the effect of immune checkpoint inhibitor (ICI) exposure on time to SJS/TEN onset. PD-1 inhibitors were significantly associated with delayed onset (HR 0.82, p < .01). Cancer diagnosis (HR 0.78, p < .001) and higher drug counts per case were also associated with longer latency. The bottom panel shows a spline-based effect of age, indicating modest delay in older patients. (B) Cox regression comparing SJS/TEN onset among cases where the primary suspect was an ICI versus a non-lCI drug. PD-1 inhibitors again showed significant latency prolongation (HR 0.71, p < .001), with similar trends seen for combination ICls. Covariate effects mirrored Panel A, supporting robust associations between ICI mechanism and delayed SJS/TEN onset.

## Discussion

Our findings confirm that ICIs are independently associated with increased risk of SJS/TEN and can synergize with high-risk small molecules to further amplify this risk. Both strong culprits (e.g., allopurinol, TMP-SMX) and weaker culprits (e.g., fluoroquinolones, macrolides) were significant predictors of SJS/TEN, but their effects were substantially magnified in the presence of ICI exposure, supporting a model of additive or supra-additive risk. Furthermore, latency analyses revealed that ICI-associated SJS/TEN presents later than non-ICI cases, with anti-PD-1 therapies showing an onset period nearly two-fold longer. These latency effects were consistent across both time-dependent and causative-agent Cox models.

Together, these results suggest a “two-hit” model, in which ICIs lower threshold for drug-specific T-cell activation. This model is supported by emerging mechanistic evidence on HLA-restricted, T-cell mediated hypersensitivity, showing that ICI can decrease threshold for T-cell activation.^6^ The delayed onset observed with ICI-associated SJS/TEN may further contribute to misattribution, increasing the risk of under-recognition or misdiagnosis in oncology settings. From a clinical perspective, our findings underscore the importance of careful co-prescribing in ICI-treated patients. These insights also reinforce the need for prospective studies and pharmacogenomic investigations to identify patients at highest risk and to develop guidelines for safer prescribing in cancer immunotherapy.

## Data Availability

All data produced in the present study are available upon reasonable request to the authors

## Data Sharing

Data and methods are available at https://github.com/capuhcheeno/SCARs_ICI-Manuscript-Scripts/tree/main/ICI%20and%20Culprit%20Analysis.

## Contributions

EMM conceived the project, wrote the manuscript, and conducted published version of all analyses. AA and DP assisted with analyses and edited the manuscript. MMP, CAS, and MSK assisted with analyses and reviewed the manuscript. EP reviewed the manuscript.

## Acknowledgements

E.J.P. is supported by the following grants from the National Institutes of Health (NIH): NIH U01AI154659, NIH P50GM115305, NIH R01HG010863, NIH R21AI139021, NIH R01AI152183, and NIH 2 D43 TW010559. E.J.P. is also supported by the National Health and Medical Research Council of Australia. E.M.M. is funded by a Vanderbilt University Medical Center internal career development award (Vanderbilt Faculty Research Scholars).

## Conflict of Interest

E.J.P. receives royalties and consulting fees from UpToDate and UpToDate Lexidrug (where she is a Drug Allergy Section Editor and section author) and has received consulting fees from Janssen, Vertex, Verve, Servier, Rapt and Esperion. E.J.P. is co-director of IIID Pty Ltd, which holds a patent for HLA-B^*^57:01 testing for abacavir hypersensitivity, and E.J.P has a patent pending for detection of HLA-A^*^32:01 in connection with diagnosing drug reaction with eosinophilia and systemic symptoms to vancomycin. For these patents she does not receive any financial remuneration, and neither are related to the submitted work.

## Statement on Use of Artificial Intelligence

GPT4o was used to write, optimize, and debug R, SQL and Python code. Suggestions were cross-referenced in literature, package documentation, and resources like stackoverflow.com. All code and generated data were manually reviewed, tested, and verified by the authors. The authors take responsibility for all code generated. GPT4o was also used for idea generation and assistance with clarity in manuscript writing.

## Data Preparation

We analyzed FAERS reports from 2013 to 2023 using a deduplicated dataset constructed as previously described.^1^ We restricted analyses to the post-ICI era (2013 onward) and excluded duplicate reports, following standard pharmacovigilance practices.

Drugs coded as primary suspect (PS) were considered causative for the sake of this analysis. TMP-SMX exposure was comprehensively captured by merging reports listing trimethoprim, sulfamethoxazole, or pre-combined formulations. Exposure variables were constructed for immune checkpoint inhibitors (ICIs) when present in the drug list for a patient in any role. ICIs were grouped by mechanistic class (PD-1, PD-L1, CTLA-4, LAG-3), and additional variables included polypharmacy (number of unique drugs), sex, age, and cancer diagnosis (assigned using both indication keywords and manual curation of cancer-specific medications).

Age was missing in 43.9% of entries. To preserve modeling power while respecting variable distribution, we imputed missing ages using sequential hot deck imputation via the impute_shd() function from the simputation package, using the ten most age-correlated variables as donors. This approach preserves covariate relationships without assuming distributional form.

For time-to-event analyses, we calculated latency (TTE, in days) as the interval between drug initiation and SJS/TEN symptom onset, using FDA-submitted dates. We retained events with full month, day, and year information for both primary suspect drug and event, with plausible latency values between 1 and 180 days. In total, 4,086 SJS/TEN cases had usable TTE data and were included in survival analyses.

## Logistic Regression Analysis

To assess whether immune checkpoint inhibitor (ICI) exposure modifies the association between small-molecule drugs and risk of SJS/TEN, we conducted a multivariable logistic regression that included interaction terms between ICI exposure and two drug classes: strong culprits (e.g., allopurinol, TMP-SMX) and weak culprits (e.g., macrolides, fluoroquinolones). The final model was specified as:

\text{logit}\left(P(\text{SJS/TEN})\right) = \beta_0 + \beta_1 \cdot \text{ICI} + \beta_2 \cdot \text{StrongCulprit} + \beta_3 \cdot (\text{ICI} \times \text{StrongCulprit}) + \beta_4 \cdot \text{WeakCulprit} + \beta_5 \cdot (\text{ICI} \times \text{WeakCulprit}) + f(\text{age}) + \beta_6 \cdot \text{sex} + \beta_7 \cdot \text{cancer} + \beta_8 \cdot \text{NumDrugs} + \beta_9 \cdot \text{ageMissing}

Where:

- f(age) is modeled using a natural spline with four degrees of freedom to capture nonlinear age effects.
- ageMissing is a binary variable indicating imputed vs. observed age, included to account for missing age values (imputed using sequential hot-deck imputation).
- Additional covariates included sex, cancer diagnosis, and polypharmacy (number of drugs).

We estimated multiplicative interaction effects for both ICI × Strong Culprit and ICI × Weak Culprit using interaction terms in the model. To assess additive interaction, we used the interactionR package to compute^2^:

- RERI (Relative Excess Risk due to Interaction) \text{RERI} = RR_{11} - RR_{10} - RR_{01} + 1
where RR_{ij} denotes risk ratios for combinations of ICI (i) and culprit exposure (j).
- AP (Attributable Proportion due to interaction) \text{AP} = \frac{\text{RERI}}{RR_{11}}
- S (Synergy Index) \text{S} = \frac{RR_{11} - 1}{(RR_{10} - 1) + (RR_{01} - 1)}

These metrics quantify the degree of risk amplification due to combined ICI and culprit drug exposure, beyond the sum of their individual effects. Confidence intervals for RERI, AP, and S were derived using delta method standard errors as implemented in interactionR.

## Time-to-Event Analyses

We performed two complementary Cox proportional hazards models to assess latency patterns in SJS/TEN cases, using documented time-to-onset (TTE) from drug initiation to symptom onset.

### Model 1: Time-Dependent ICI Exposure

To address immortal time bias and examine mechanistic differences across immune checkpoint inhibitor (ICI) classes, we implemented a time-dependent Cox regression using interval-split data. Each patient’s follow-up time was divided into risk intervals corresponding to exposure status for PD-1, PD-L1, CTLA-4, and LAG-3 inhibitors before the event date. For those cases with only month or year information, admin date was coded as the beginning of the month or the year (Jan 1), respectively. Time-dependent binary indicators were updated dynamically at the start of each interval to reflect ICI administration timing to avoid immortal time bias. The model was fit using the following structure:

\lambda(t) = \lambda_0(t) \exp\left(\beta_1 \cdot \text{PD-1}_t + \beta_2 \cdot \text{PD-L1}_t + \beta_3 \cdot \text{CTLA-4}_t + \beta_4 \cdot \text{LAG-3}_t + f(\text{age}) + \boldsymbol{\beta} \cdot X\right)

Where:

- \(\lambda(t) \): Hazard function at time \(t \)
- \(\lambda_0(t) \): Baseline hazard at time \(t \)
- \(\text{PD-1}_t, \text{PD-L1}_t, \text{CTLA-4}_t, \text{LAG-3}_t \): Time-dependent ICI exposure indicators
- \(f(\text{age}) \): Penalized spline function for age (4 degrees of freedom)
- \(\boldsymbol{\beta} \cdot X \): Linear predictor for covariates (e.g., sex, cancer status, region, number of drugs)

This approach allowed for estimation of hazard ratios specific to ICI subclasses and identification of delayed versus early-onset risk patterns associated with different immunotherapy agents.

### Model 2: ICI vs Non-ICI Culprit Comparison

To compare latency distributions between ICI-attributed and non-ICI-attributed SJS/TEN cases, we restricted analysis to reports with clearly identified primary suspect drugs. We classified each causative agent as an ICI or non-ICI and further subtyped ICIs by mechanism (PD-1, PD-L1, CTLA-4, or combinations thereof). A standard Cox proportional hazards model was then constructed using TTE as the outcome:

λ(t)=λ0(t)ex (β⋅Mechanism+f(age)+β⋅X)\lambda(t) = \lambda_0(t) \exp\left(\beta \cdot \text{Mechanism} + f(\text{age}) + \beta \cdot X\right)

As above, covariates included penalized splines for age, along with sex, cancer status, polypharmacy, and region. Mechanism of action (e.g., PD-1, PD-L1, CTLA-4, CTLA-4/PD-1) was treated as a categorical variable with “non-ICI” as the reference group. ICIs administered in combination were collapsed into a single composite variable using the earliest shared start date and unique mechanism combinations (e.g., “CTLA-4 / PD-1”).

## Visualization and Interpretation

For logistic regressions, marginal effect plots for age and interaction terms were generated using the ggpredict R package. For Cox models, hazard ratios with 95% CI were extracted from each model and visualized via forest plots. Age effects were displayed separately using spline-predicted hazard functions, plotted against binned age values with ribbons representing ±1.96 standard error. Spline terms captured nonlinear associations between age and latency. All models were implemented in R using the survival, pspline, and broom packages.

## Notes

### Author Declarations

FDA FAERS database

### Summary of Updates

1. Age has been incorporated into all models as a continuous variable using spline terms. Specifically, we used natural splines with 4 degrees of freedom for logistic regression and penalized splines for Cox models, as recommended. 2. We revised our approach to focus on a single comprehensive logistic regression model including interaction terms between ICI exposure and both strong and weak culprit drugs. We additionally report additive interaction metrics-including RERI, attributable proportion (AP), and synergy index (S) calculated using the `interactR` package. 3. We replaced the unadjusted Kaplan-Meier plot with two multivariable Cox models presented as forest plots (Figure 2), showing adjusted hazard ratios with N and number of events. Latency was analyzed using both time-dependent exposure models and models based on causative drug class. 4. To address concerns about immortal time bias, we implemented a time-dependent Cox model in which ICI exposure status was updated dynamically based on reported drug start dates relative to the index event.

